# Utilization of Generative AI-drafted Responses for Managing Patient-Provider Communication

**DOI:** 10.1101/2025.08.31.25334725

**Authors:** Soumik Mandal, Batia M. Wiesenfeld, Adam C. Szerencsy, William R. Small, Vincent Major, Safiya Richardson, Antoinette Schoenthaler, Devin Mann, Oded Nov

## Abstract

The integration of generative AI (GenAI) in patient communication presents benefits and challenges. This retrospective observational study analyzed EHR audit logs to assess how 75 healthcare professionals (HCPs) utilized AI-generated drafts for patient messages from October 2023 to August 2024 at a large health system in New York City. Overall utilization was low (19.4%), though prompt refinements improved usage (from 12% to 20%), particularly among physicians. GenAI drafts were generated for all messages, including 80% that received no response, adding to the review burden and potentially undermining efficiency. Text analysis showed HCPs preferred concise, information-rich drafts, with role-based differences— physicians favored shorter drafts, while clinical support staff preferred more empathetic responses. AI-generated drafts reduced message turnaround time by 6.76% despite a marginal increase in required steps (*InBasket actions*). These findings highlight the need for targeted GenAI deployment strategies, better aligned with clinician workflows and optimized draft generation for improved efficiency.

## Introduction

The increasing volume of patient messages in electronic health record (EHR) inboxes reflects a broader shift toward asynchronous, digital care that may be more accessible and efficient for patients.^1^ Without dedicated time or workflow support, healthcare professionals (HCPs) are often left to manage this demand unsustainably, adding to their administrative burden,^2,3^ worsening burnout.^4–6^ In response, healthcare systems have implemented various inbox management strategies^7^ aimed at supporting HCP workload while maintaining patient communication quality. These include filtering low-clinical-value messages, delegating messages to Advanced Practice Providers (APPs) and support staff,^8^ automating message triage,^9^ and implementing billing policies to reduce avoidable patient messages.^5^ However, these interventions have costs and their effectiveness is limited,^4^ highlighting the need for more efficient and scalable solutions to manage the growing patient message volume. To address this, health systems are increasingly deploying large language model (LLM)-based generative artificial intelligence (GenAI)^10^ to draft message replies.^11–15^ However, its impact on reducing HCP’s efforts to address patient messages remains unclear,^14,16^ and little is known about how these drafts affect response quality and timeliness or affect patient outcomes.^17,18^

GenAI can accurately apply medical knowledge,^19,20^ and generate contextually relevant responses.^21,22^ Studies evaluating GenAI-drafted responses to patient messages have found them comparable to human responses in acceptability, accuracy, and empathy by both patients and providers.^14,23–26^ When integrated into the EHR inbox (referred to as the InBasket in our EHR and hereafter in this paper), GenAI can leverage patient-specific information from the EHR and draft responses to patient messages that HCPs can review and edit before sending to patients.^12,27^ This approach can help alleviate the cognitive load associated with responding to patient messages,^28^ which often requires HCPs to navigate through interfaces, search the EHR for relevant information and compose responses while managing other complex clinical tasks, and places competing demands on their working memory.^29^ This process is inefficient due to navigational challenges in EHR, poor interface design, and usability issues including excessive clicks, limited customization (e.g., inability to hide rarely used folders), unclear icons, and confusing layouts.^30^ A recent qualitative study across six health systems identified over 50 EHR-related barriers to efficient message processing.^31^ While reviewing GenAI-generated drafts may require additional time than reviewing patient messages alone,^14,32^ it is unknown whether the time spent is offset by reductions in task-switching and information retrieval from EHR.^33^ Although GenAI has the ability to extract and summarize relevant EHR information,^34^ current implementations fall short of this promise.^35^ Ultimately, the value of GenAI augmentation^36^ within the InBasket depends on whether and how it streamlines patient communication, improves provider efficiency, and enhances patient outcomes.^37^

Despite its potential, GenAI adoption in patient communication remains largely experimental, with most institutions piloting but not fully embracing it.^38^ Prior evaluations have provided inadequate evidence of impact on provider efficiency, with many pilot studies reporting low utilization rates by providers.^11,28^ This limited uptake reflects broader skepticism toward digital health technologies,^39^ driven by concerns about accuracy, liability, reduced autonomy, depersonalized patient interactions, and AI-generated hallucinations (fabricated information that could mislead patients),^40,41^ as well as apprehensions that such tools prioritize institutional efficiency over patient care.^14^ Key limitations of prior evaluations include reliance on simulations and surveys measuring perceived rather than actual utilization, reducing real-world applicability.^15,24^ Studies reporting utilization at the provider-level rather than message-level^11,28^ risks biasing usage estimates.^15^ Most evaluations emphasize time-based metrics^12^ while overlooking the complexity of managing patient messages—such as reading inquiries, reviewing charts, and interpreting lab results^42,43^—tasks that are known contributors to task-switching and cognitive load.^44,45^ EHR audit logs, which provide detailed records of providers’ actions,^43,46–48^ can be leveraged for evaluating the utility of GenAI-drafted responses to patient messages,^37^ providing real-world insights into their impact on clinical workflow and efficiency.

This study attempts to address these critical gaps by examining trends in the utilization of GenAI-drafted responses to patient messages over time and across different user roles, and how quality-relevant features of generated drafts influence utilization. It uses EHR audit log data to compare the steps taken by HCPs in managing patient messages, evaluating the impact of using generated drafts on workflow. By identifying factors that facilitate or hinder utilization, this research aims to inform strategies for optimizing GenAI implementation for managing patient messages in clinical practice. The Methods section details the study setting, the clinical sites that piloted GenAI drafts for patient messages, GenAI integration, the participating users and their roles in healthcare delivery, as well as the types of patient messages and corresponding provider activities analyzed.

## Results

### Sample Description

Of the 75 pilot HCPs included in the study, 54 were physicians (72%), 14 were clinical support staff (18.67%), and 7 were administrative support (9.33%). Between October 11, 2023 (when the last pilot HCP started receiving GenAI-drafted responses) and August 31, 2024, a total of 55,767 unique patient messages were received that were either addressed to or responded by the HCPs. Approximately 80% (44,454) of these messages (e.g., thank you notes, consecutive patient messages prior to HCP’s response) were resolved without any response sent to the patient by the HCPs. For these 44,454 messages, length of GenAI-generated drafts averaged 56 words per message (min: 3, max: 330, SD: 27.56, 95% CI: 55.31 to 55.83). Based on an average adult silent reading rate of 238 words per minute (wpm),^49^ the estimated mean reading time for these drafts is 13.51 seconds (min: 0.76 seconds, max: 83.19 seconds, SD: 6.95 seconds, 95% CI: 13.94 seconds to 14.08 seconds) per message, totaling approximately 10,006.29 HCP-minutes relative to 7,389.08 HCP-minutes for reading the patient messages themselves. Of the remaining 11,313 patient messages, drafts were displayed in at least one pilot HCP’s InBasket in 5,935 instances (52.5%) and were used as the denominator for calculating overall utilization.

### GenAI Draft Utilization

In 1,149 of the 5,935 eligible instances, HCPs read a patient message and used the “Start with Draft” option to compose a response (see **Supplementary Figure 1**), yielding a utilization rate of 19.4%. Only 3.3% (n = 196) of messages were initiated using the “Start Blank Reply” function. Utilization improved significantly over time (F(8,126) = 2.68, p = 0.009) increasing from period 1 with the out-of-the-box prompt (mean= 0.12, SD=0.13, 95% CI=0.09–0.18) to its peak in the period 3 (see **Supplementary Figure 2**) following the February 28, 2024 super-prompt update (mean=0.20, SD=0.14, 95% CI=0.14–0.22). HCP type was an important predictor of draft utilization (see **Figure 1**). The administrative support group consistently showed the highest utilization across all periods, peaking in period 3 (mean=0.32, SD=0.12, 95% CI=0.27– 0.37). Physicians’ utilization was at its highest during the final period (mean=0.12, SD=0.05, 95% CI=0.10–0.14), while clinical support staff demonstrated their greatest utilization in period 1 (mean=0.14, SD=0.08, 95% CI=0.07–0.20) (see **Supplementary Table 4**). Comparison of utilization across the three periods showed significant differences for physicians (test-stat = 23.74, *p* <0.001), but not for the other two groups.

**Figure 1:**
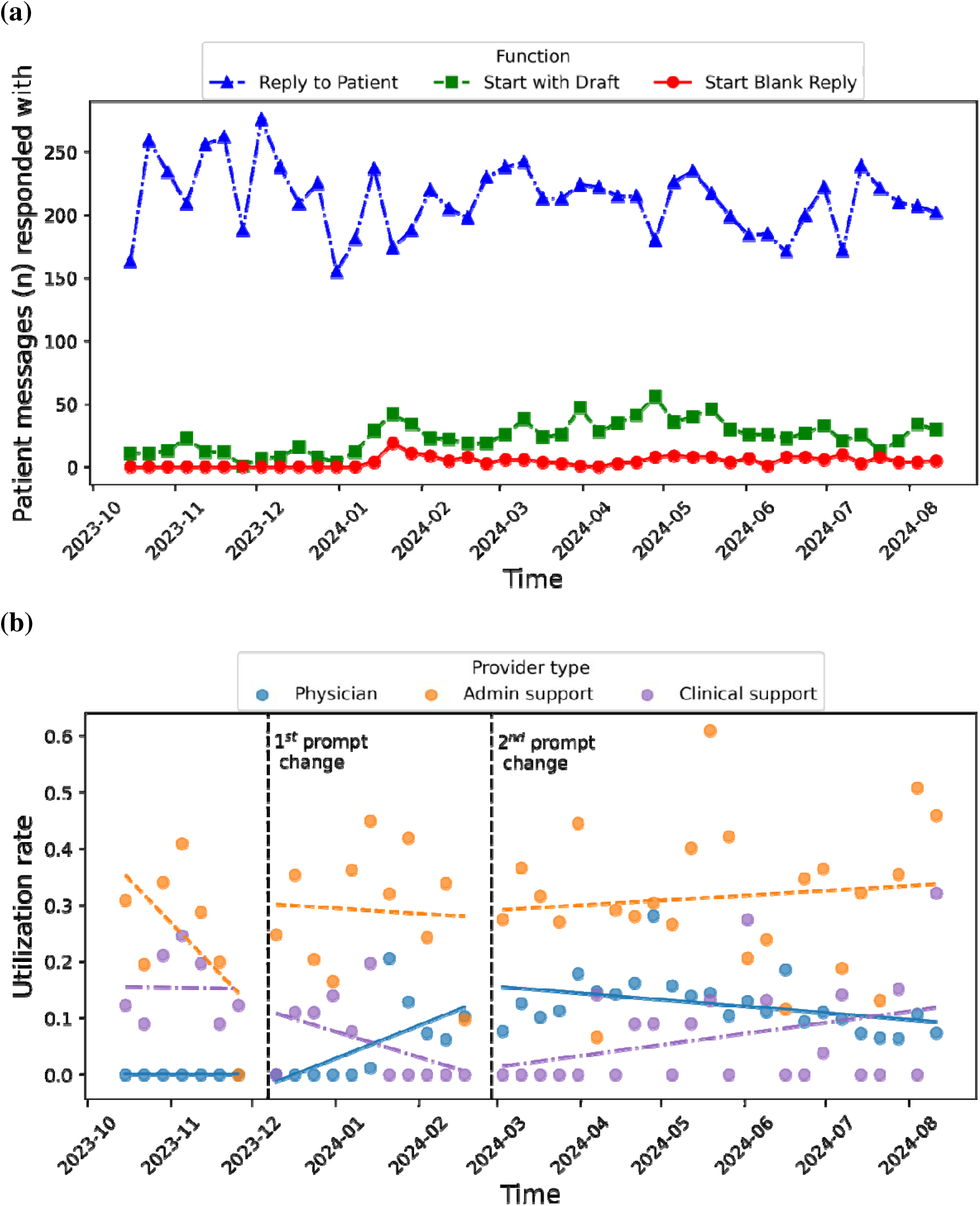
Trends in utilization: (a) temporal utilization of all three functions to reply to patient messages across all HCPs; (b) temporal utilization of GenAI drafted response with linear trend lines compared between HCP types.

### Draft Characteristics Influencing Utilization

The logistic regression model identified several draft characteristics (Table 2) as significant predictors of draft utilization. Among characteristics derived from the draft alone, informativeness (i.e. entropy), sentiment subjectivity, reading ease, affective content and brevity were significant predictors of utilization (see **Figure 2**). Specifically, entropy (β = 0.4047, *p* < 0.001, 95% CI: 0.125 to 0.685), sentiment subjectivity (β = 0.4050, *p* = 0.010, 95% CI: 0.098 to 0.711) and Flesch reading score (β = 0.0161, *p* < 0.001, 95% CI: 0.011 to 0.022) were all positively associated with utilization. Mean sentence length also showed a positive association with utilization (β = 0.0549, p = 0.002), but word count was negatively associated with utilization (β = -0.0108, p < 0.001, 95% CI: -0.017 to -0.005), suggesting that users preferred drafts with fewer, but more informative sentences. Relating draft characteristics to patient messages, semantic similarity with patient messages was positively linked with utilization (β = 1.2968, *p* < 0.001, 95% CI: 0.759 to 1.835), while content overlap with patient messages had a small but significant negative effect (β = -0.3032, *p* = 0.028, 95% CI: -0.574 to –0.033).

**Figure 2:**
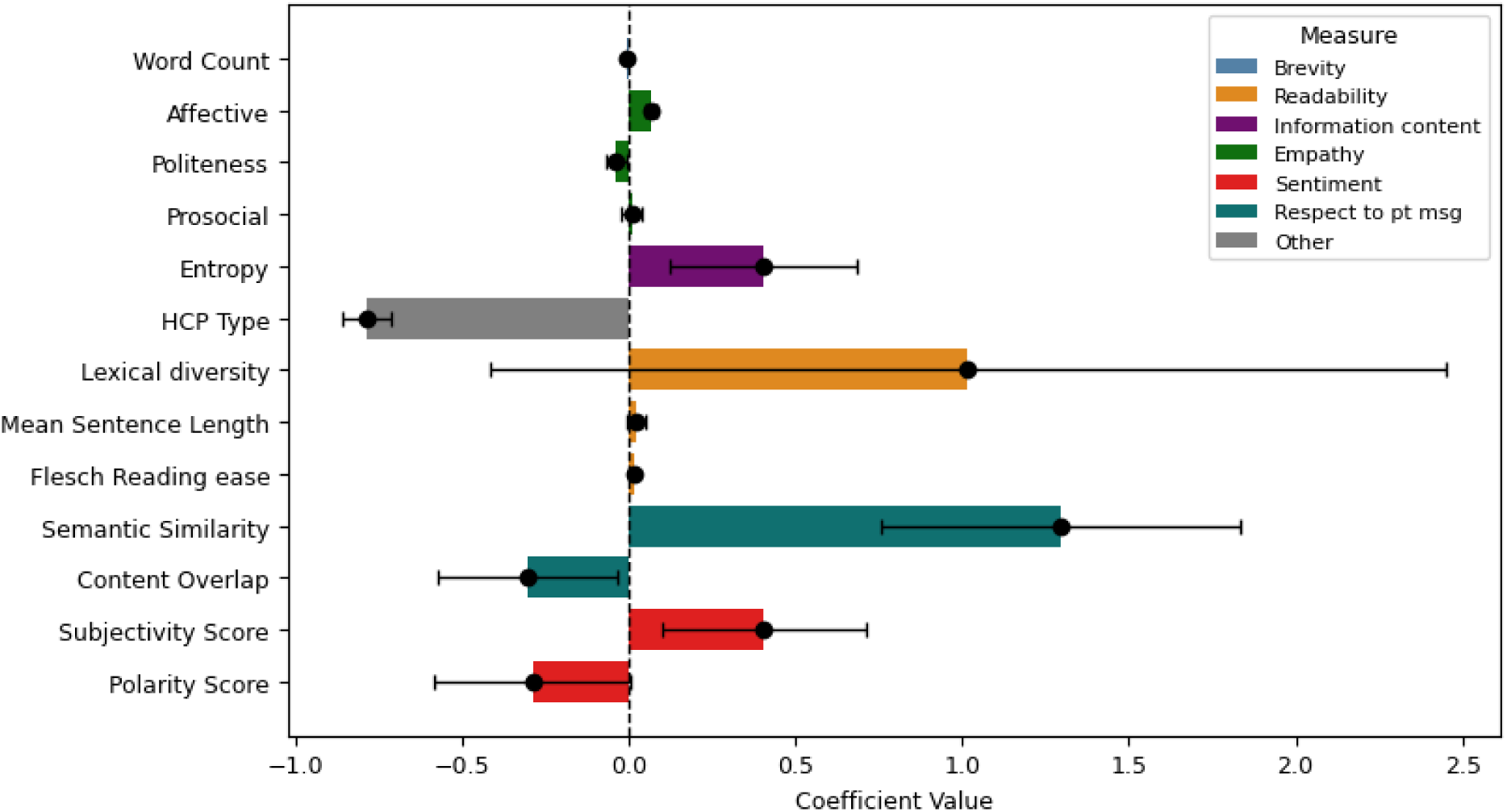
Predicted utilization of GenAI-generated drafts by healthcare professionals (HCPs) based on linguistic features. Positive coefficients indicate features that increase utilization while negative coefficients reduce.

The importance of draft characteristics as determinants of the likelihood of draft utilization varied across HCP types. Post-hoc tests revealed that physicians were less likely to use longer drafts, utilizing those with fewer words than clinical support (mean difference = -9.70, *p* < 0.001) and administrative support (mean difference = -8.15, *p* < 0.001). Drafts utilized by physicians also had lower readability, evident by lower Flesch reading ease scores than those used by administrative support (mean difference = -1.99, *p* < 0.001) and clinical support (mean difference = -2.43, *p* < 0.001). In contrast, clinical support staff favored drafts with higher affective content than physicians (mean difference = 0.7216, *p* < 0.001) and administrative support (mean difference = 0.4419, *p* = 0.012), with similar differences also observed for sentiment subjectivity, entropy scores.

Semantic similarity to patient messages also varied by HCP type, with drafts used by administrative support showing lower semantic similarity to patient messages than those used by physicians (mean difference = -0.0189, *p* < 0.001) and clinical support (mean difference = -0.0235, *p* = 0.008), and lower content overlap with patient messages compared to physicians (mean difference = -0.0264, *p* < 0.001). These results are consistent with those expected if administrative support users focused their responses more on administrative issues and less on addressing patients’ medical concerns.

### Patient and Message Characteristics Influencing Utilization

We also explored whether draft utilization was influenced by factors on the patient side. Patient complexity did not influence draft utilization; after adjusting for age, the mean Charlson Comorbidity Index (CCI) of patients was 2.22, with no significant difference between groups (difference: -0.13, *p* = 0.66). To assess whether draft utilization varied depending on the cognitive load required to understand the patient message, we evaluated message complexity and found that message complexity was associated with increased GenAI draft utilization. Messages responded to with GenAI drafts were less readable than those without, as reflected by a lower Flesch reading score (difference: -1.27, *p* = 0.005) and a higher syllables-per-word count (difference: 0.02, *p* = 0.01). No significant differences were observed in lexical diversity or sentence length (see **Supplementary Table 5**).

Overall, these findings show that draft characteristics, particularly information content, relevance and completeness are key factors influencing the likelihood of message utilization, but the importance of these factors varied across HCP types. They also suggest that HCPs used GenAI drafts when the messages patients sent were more difficult to understand with respect to readability.

### Efficiency Evaluation

HCPs responded to 11,313 patient messages, generating 52,397 InBasket actions, with an average of 4.65 actions per message (range: 2–113; SD: 3.4; 95% CI: 4.59-4.72). A subset (7.41%, *n* = 838) of patient messages were missing second-level timestamps in action logs, leaving 10,475 messages for time analysis. The median turnaround time was 354 seconds (IQR: 14,264 seconds). The median message open time was 55 seconds (IQR: 5,097 seconds) and the median time since last opened was 17 seconds (IQR: 61 seconds). Among the 20 distinct action types recorded **(Supplementary Table 3**), **view reports**, which involve opening the message without any additional steps taken, accounted for 56.4% (n = 29,540) of all actions. On average, each message required 2.76 unique actions (range: 2–7; SD: 0.8; 95% CI: 2.75–2.78) and 1.89 repeated actions (range: 0–106; SD: 3.01; 95% CI: 1.83–1.94). Additionally, messages were forwarded to another HCP in 669 instances, approximately 1 in every 17 messages.

First, we evaluated potential time-related efficiency gains (see **Table 2**) from GenAI draft utilization by comparing time metrics between messages responded to with and without GenAI drafts (see **Figure 3**). Responses utilizing GenAI drafts had a 6.76% shorter median turnaround time than those that did not utilize GenAI drafts (median [utilized]: 331 seconds, median [not utilized]: 355 seconds, difference: -24 seconds, *p* < 0.001). However, the median message open time increased by 7.27% (median [utilized]: 59 seconds, median [not utilized]: 55 seconds, difference: 4 seconds, *p* < 0.001), and the median time since last opened (unattended time) increased by 15% (median [utilized]: 23 seconds, median [not utilized]: 20 seconds, difference: 3 seconds, *p* < 0.001). (See **Supplementary Table 6** for additional details).

**Figure 3:**
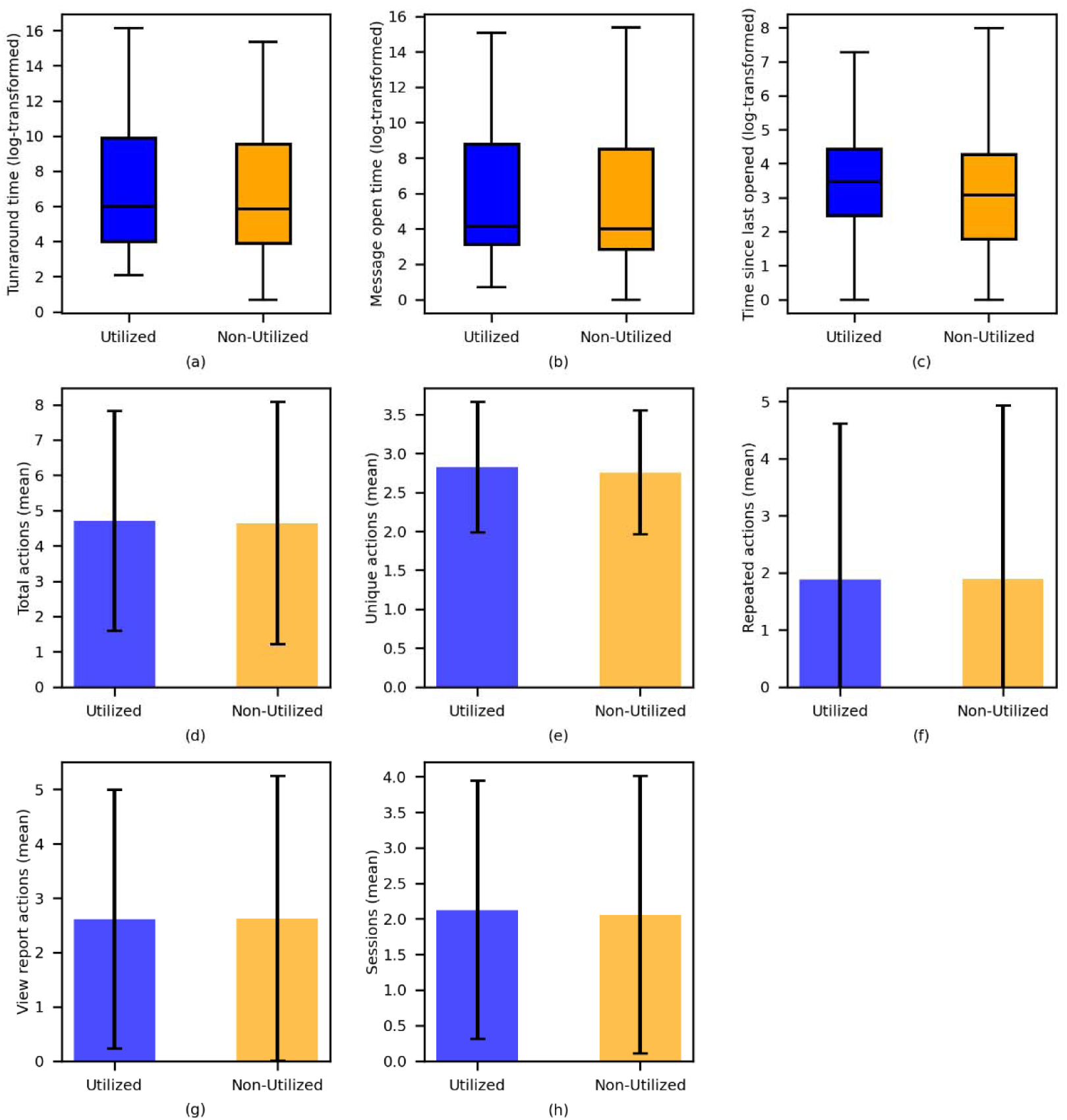
Comparison of GenAI-draft utilized and non-utilized messages. Box plots and bar charts (with SD as error bars) show distribution of time-related measures and InBasket actions, with the utilized group in blue and non-utilized in orange. Subfigures include: **(a)** turnaround time, **(b)** message open time, **(c)** time since last opened, (**d**) total actions, (**e**) unique actions, **(f)** repeated actions, (**g**) view report actions, and **(h)** session counts. Time-related measures (**a**, **b**, **c**) were log-transformed to account for skewness and reduce the influence of extreme values in the figure.

Given that faster response time when utilizing GenAI drafts could not be explained by earlier message opening or reduced unattended time, we analyzed InBasket actions to understand the source of time efficiency gains. Comparing all InBasket actions for messages in which GenAI drafts were utilized to those in which they were not utilized revealed that HCPs took slightly (1.95%) more actions (mean [utilized]: 4.71, mean [not utilized]: 4.62, difference: 0.09, *p* = 0.38) per message when utilizing GenAI drafts (see **Supplementary Table 7**). When removing the reply actions (*Reply to patient*, *Message Patient*, and *IB suggested response*), which differed for GenAI drafts, the difference was even smaller (mean [utilized]: 2.86, mean [not utilized]: 2.84, difference: 0.02, *p* = 0.45). GenAI draft utilization was associated with an increase in unique actions (mean [utilized]: 2.82, mean [not utilized]: 2.76, mean difference: 0.06, *p* = 0.02), while repeated actions (mean [utilized]: 1.88, mean [not utilized]: 1.89, mean difference: -0.01, p = 0.94) and message views (mean [utilized]: 2.61, mean [not utilized]: 2.62, mean difference: -0.01, p = 0.73) remained nearly unchanged (see **Supplementary Table 7**).

While the number of actions differed relatively little, the frequency of a few types of actions differed when responses utilized the GenAI draft (see **Supplementary Table 8**). In particular, there were fewer shifts of responsibility (any of the following actions: *Take responsibility*, *Take put back responsibility submenu, Put responsibility back,* and *Move to My Messages*) for responses that utilized GenAI drafts (mean [utilized]: 0.03, mean [not utilized]: 0.07, difference: 0.04, *p* = 0.04) but more create telephone call (mean [utilized]: 0.03, mean [not utilized]: 0.06, difference: -0.03, *p* = 0.01), and encounter for medical review actions (mean [utilized]: 0.04, mean [not utilized]: 0.06, difference: -0.02, *p* = 0.07). Additionally, no significant association was observed between draft utilization and message forwarding among HCPs (χ² = 0.04; *p* = 0.84; see **Supplementary Table 9** for details).

## Discussion

In this comprehensive evaluation using electronic health record (EHR) audit logs, we examined how healthcare professionals (HCPs) utilized these drafts in patient communication, assessing differences in utilization across roles, the characteristics of drafts that increase utilization, and the impact of utilization on providers’ efficiency and workload. Our analysis shows overall utilization improved over time but remained low at 19.4%. Our study spanned 11 months, far longer than prior institutional pilots lasting only a few weeks,^11^ yet utilization rates remained as low as early GenAI deployments,^9^ and far lower than the rates implied by experimental findings.^24^ Prompt refinements have significantly improved utilization over time, particularly among physicians.

We applied a novel text-analysis framework to understand how draft content affects use rather than benchmark the underlying large language model (LLM),^50^ which has been extensively evaluated elsewhere.^51^ Our analysis of the relationship between draft content and utilization rate suggests that drafts with more informative content, greater subjective language and clearer readability were more likely to be utilized. Drafts with greater relevance to the patient message were used more often, but those with higher overlap of content from the patient message were used slightly less. Overall, these findings suggest that drafts likely to serve patient needs better were more likely to be utilized. Some improvement on these dimensions may be achieved with better prompts while further improvement may require fine-tuning the LLM itself but understanding what characteristics most strongly influence utilization enables more efficient progress toward implementation objectives.

Overall draft utilization rates varied by role with administrative support staff consistently demonstrating higher utilization, regardless of prompt updates. These findings mirror those in other sectors that have demonstrated GenAI’s ability to bridge expertise gaps for lower-skilled workers.^52^ If similar benefits are realized in patient communication, it could enhance efficiency by enabling support staff—who typically handle fewer messages^53^— to manage a larger share of routine patient interactions, particularly those that do not require input from physicians or specialists. By improving communication quality, GenAI can catalyze the professional development of support staff and enhance patient care.

The draft qualities that most strongly influenced utilization differed across HCP roles, suggesting that implementation success may depend on customizing the technology to user role. Physicians preferred shorter yet more complete drafts, while clinical support staff utilized drafts with more affective content, suggesting that they prioritized expressing empathy in patient interactions.^54^ Administrative support staff, in contrast, utilized drafts less relevant to patient messages, which can be expected due to their non-clinical advisory role. These role-based differences emphasize the need for AI-generated drafts to be tailored not only for content quality but also for appropriate use across HCP groups.^15^

Overall, draft utilization was greater when the original patient message was less readable. Prior work has shown that InBasket management is time-consuming, burdensome and burnout-inducing.^55^ More complex and difficult to understand patient messages exacerbate cognitive load. Team-based care interventions designed to aid InBasket management redirect less complex messages to non-physician HCP’s, but do so without alleviating the cognitive load of complex patient messages.^8,55^ The inverse relationship between reading complexity and utilization may reflect demand for GenAI that succinctly summarizes patient messages in accessible language that requires less effort to reply to, but this could potentially raise safety concerns.

The burnout and burden associated with InBasket work has sparked interest in GenAI solutions that can increase HCP’s InBasket management efficiency. HCPs are familiar with technologies portrayed as increasing efficiency that in practice impose new and greater time demands that sap efficiency.^56^ Our analysis identified such a risk in the need to review unnecessary drafts, which were generated and displayed for all patient messages, without considering whether a response is necessary. While it is difficult to determine with certainty whether HCPs read drafts in such cases, our estimates suggest that when drafts are reviewed, they add a significant burden, increasing the time spent on each message by 135.42% compared to reading the patient message alone. Recent perspectives in digital health implementation increasingly emphasize digital minimalism, advocating for a deliberate, optimized approach to avoid overwhelming clinicians.^57^ The core tenets of digital minimalism—“*clutter is costly, optimization is vital, and intentionality is essential*”—are particularly relevant here.^58^ Even if unnecessary drafts are not always read, their presence adds to InBasket clutter, a known stressor linked to physician burnout.^59^ Prior work in implementation science has highlighted the importance of workflow integration in determining the success of health technologies; poorly integrated tools often fail due to misalignment with end-user needs.^60^ A more targeted AI deployment strategy, focused on high-yield patient message types, can improve acceptance and utilization among HCPs.

Our analysis of message actions and time metrics revealed that AI-generated drafts significantly impact patient message handling. We found evidence that messages completed without drafts required 6.76% more time to complete despite involving similar numbers of actions, suggesting that utilization may streamline message management. These findings are consistent with prior studies that showed, despite AI-generated replies leading to increased read time,^12^ healthcare providers experience lower cognitive load when *editing* AI-generated drafts for patient responses, compared to writing replies from scratch.^13^ Our data was collected over a longer time period post-implementation when users may have been farther along the learning curve. Notably, message forwarding rates were not significantly associated with draft utilization, indicating that GenAI did not enable support staff to independently handle more patient messages. This suggests that while the drafts improve individual message processing, their impact on broader workflow efficiency remains limited in the current implementation, representing an important area for further analysis and improvement. There is an urgent need to improve draft utilization before GenAI can meaningfully reduce the InBasket burden of patient messages on HCPs.

More fine-grained analysis of actions offers insight into how message handling may change with GenAI draft utilization. Messages completed without drafts involved more shifts of responsibility. On the other hand, those in which drafts were utilized were more likely to involve creating a telephone call, which itself imposes additional time requirements. These results are also consistent with broader research on human-centered AI that suggest automated systems can streamline workflows but may also introduce new complexities or time burdens.^61^

This study has several limitations. It was conducted within a single health system, and findings may not reflect experiences in other settings with different workflows, staffing models, or patient populations. The evaluation did not include qualitative feedback from HCPs to explain why drafts were or were not used. Additionally, the absence of human annotation and structured review of a random sample of drafts limits insights into draft quality and end-user perceptions.

Comparisons across HCP types may be biased due to class imbalance, with physicians comprising the majority of users. Temporal factors such as holiday schedules may have further influenced message volume and draft utilization. Despite these constraints, our findings provide critical insights into how GenAI-generated drafts impact healthcare communication, highlighting both efficiency gains and future implementation challenges. Broader adoption will require optimization of AI-generated responses to enhance usability and better align with provider needs. Future research should incorporate multi-site, mixed-methods evaluations to capture usage patterns and end-user perspectives, refine draft generation strategies, assess patient perceptions, and ensure that AI integration supports clinical workflows without introducing unintended burdens.

## Methods

### Settings, and Gen AI integration

This prospective observational quality improvement study was conducted at New York University Langone Health (NYULH), a large academic health system in New York City. A total of 108 HCPs including physicians and their support staff from three clinical sites (two Internal Medicine and one Neurology department) participated in a phased rollout. Prior to using the generated drafts, HCPs were required to accept a one-time disclaimer affirming their understanding and agreement to use the tool responsibly. Only those who completed this step (n=75) were included in the study; HCPs who did not accept the disclaimer at the time of data collection (n=33) were excluded from all analyses. Messages designated as non-urgent medical advice requests by patients at submission were routed to the EHR vendor (Epic Systems) upon inbox delivery, which automatically classified them into four predefined groups: *general inquiries*, *test results*, *medications*, and *paperwork*. Initially, this classification triggered a separate prompt for each category, leveraged patient-specific information from the EHR (e.g., the patient’s problem list, medication list, recent and upcoming appointments) to provide additional context to the message. Later in the study, the process shifted to a unified super prompt, which was used for all message types (see **Supplementary Table 1**). During the study period, NYULH implemented two rounds of iterative prompt improvement based on initial user feedback, transitioning from an initial *out-of-the box* prompt from Epic to a more refined prompt that included instructions more specific to the health system’s existing processes (see **Supplementary Table 1**). GenAI-drafted responses were then seamlessly embedded within the InBasket clinical messaging interface. When HCPs opened a patient message in their InBasket, an AI-generated draft response was displayed alongside the original message, with three response options presented simultaneously on the same screen: (a) *“Start with the draft”*—use the draft as a starting point, edit as needed, and send; (b) “*Start blank reply*”—compose response from a blank screen; or (c) use the existing “*Reply to patient*” function to write a message independently (see **Supplementary Figure 1**). Options (b) and (c), functionally redundant in the system design, both allowed HCPs to compose responses from blank screen and were classified as non-utilization of the AI-generated draft.

### Data Collection and Measurement

HCPs were classified into three groups based on their service-role: physicians, clinical support (e.g., medical assistants, registered nurses), and administrative support (e.g., patient support associates, front desk, manager) (see **Supplementary Table 2**). The dataset included all patient-initiated messages managed by participating HCPs during the study period, for which AI-drafted responses were generated and displayed (see **Supplementary Note 1** for inclusion exclusion criteria). Each message was assigned a unique identifier by the electronic health record (EHR) system. The data collection process tracked messages using these identifiers through EHR audit logs, capturing clinical activities performed by HCPs from the message’s entry into their InBasket until it was marked as “*Done*,” signifying that no further action was needed (**Figure 4**). This ensured that all relevant actions by HCPs were accurately and chronologically linked to the corresponding messages.

**Figure 4:**
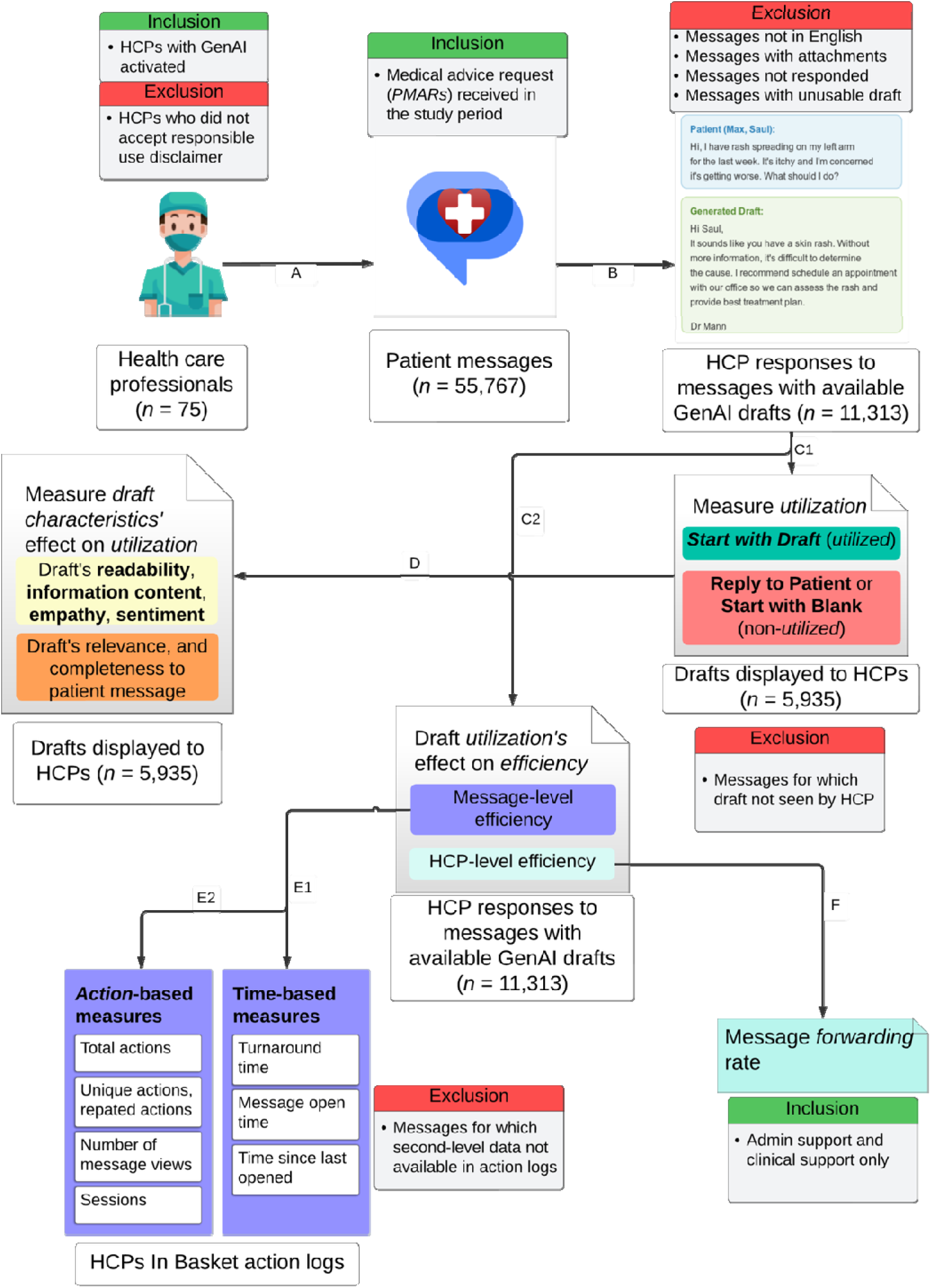
Data collection and evaluation workflow. Labeled arrows (A, B, C1, C2, etc.) denote sequential steps in the process, including data acquisition, application of inclusion/exclusion criteria, and analysis.

#### Measuring GenAI Draft Utilization

GenAI draft utilization was defined as the proportion of patient-initiated messages for which HCPs used the generated draft as a starting point for editing and sending a response. Messages were excluded from measuring utilization if they: a) did not require a response (e.g., marked “*Done*” without a response), b) had unusable drafts, such as a system error message, or c) involved generated drafts that were not displayed to any participating HCP. The latter occurred when HCPs completed messages via an alternative interface where drafts were not shown, despite being enabled in the InBasket (see **Supplementary Note 1** for inclusion exclusion criteria). To analyze trends, utilization was evaluated at two-week intervals, mitigating the potential biases of longer monthly periods—influenced by seasonal fluctuations in patient inquiries—and shorter weekly periods, which could be disproportionately affected by provider unavailability. Utilization was further stratified and compared across three time periods defined by GenAI prompt updates: period 1 with *out-of-the-box* prompt by the EHR vendor (11^th^ October 2023-6^th^ December 2023), period 2 with first prompt change (7^th^ December 2023-27^th^ February 2024), and period 3 with second prompt change (28^th^ February 2024-31^st^ August 2024). **Supplementary Table 1** summarizes prompt description, prompt changes, and user feedback on the drafts that informed these revisions (see **Supplementary Figure 2** for inclusion exclusion criteria for prompt revision timeline). The underlying LLM (GPT-4) remained unchanged throughout the study period. Utilization patterns were also analyzed across three HCP categories to identify potential variations. The GenAI implementation resulted in drafts being generated for all messages, regardless of whether a response was needed. The reading time cost of reviewing these drafts was estimated based on an average adult silent reading rate of 238 words per minute (wpm).^49^

#### Draft Linguistic Characteristics Influencing Utilization

To identify factors influencing GenAI-draft utilization, we analyzed the draft characteristics that HCPs likely consider when deciding whether to utilize them. These measures were selected to assess the quality of the generated drafts, not the performance of the language model that produced them, which remained constant throughout the study. We evaluated draft characteristics both independently and in relation to the patient messages. When analyzing drafts alone, we considered **brevity, readability, informativeness and empathy**– dimensions commonly assessed in prior evaluations of GenAI drafts and considered relevant to both HCP’s decision-making and patient experience.^14,24,62,63^ **Brevity** was assessed as draft length and was measured by the draft’s word count. **Readability** was evaluated using the Flesch reading ease score, lexical diversity, mean sentence length, and mean syllables per word. **Informativeness** was quantified using entropy, a measure of how unpredictable words in text are. Repetitive or redundant text is predictable but not informative, while higher entropy indicates text with rich information content that is generally less predictable.^64^ **Empathy** was evaluated through LIWC-22 (Linguistic Inquiry and Word Count), a widely used tool in social science that analyzes the proportion of prosocial, polite, and affective dimensions.^65^ Additionally, sentiment analysis was performed to measure subjectivity (range: 0 to 1, with higher values indicating greater subjectivity) and polarity (range: -1 to 1, with higher values reflecting a more positive tone).^66,67^

When evaluating drafts in relation to patient messages, we adopted a question-answering framework derived from the text analysis literature, treating patient messages as questions and drafts as potential answers, with draft quality assessed for **relevance** and **completeness**. **Relevance** was determined by measuring semantic similarity between the draft and the patient’s inquiry using Sentence-BERT (SBERT), a widely used transformer-based model that produces sentence embeddings for efficient comparison.^68,69^ **Completeness** was evaluated by assessing content overlap, where key terms in patient messages were identified using named entity recognition (NER) and cross-verified against the drafts, following established approaches for clinical information extraction.^70,71^ The SpaCy library, which provides pretrained NER models, was used for entity extraction. Details on these measures are provided in **Table 1**.

**Table 1:** Evaluation Metrics for Linguistic Characteristics of Generated Drafts.

| Evaluation Type | Measure | Measured As | Description |
| --- | --- | --- | --- |
| Drafts Alone | Brevity | Word counts | Measured by the total number of words in the draft. |
|  | Readability | Flesch Reading Ease score | A readability score assessing how easy the drafts are to read based on sentence length and syllable count. |
|  |  | Mean sentence length | Average number of words per sentence in the draft. |
|  |  | Mean syllables per word | Average number of syllables per word in the draft. |
|  |  | Lexical diversity | Ratio of unique words to total words in the draft. |
|  | Information Content | Entropy | Measured by assessing how evenly words are distributed; low entropy means less information present. |
|  | Empathy | Prosocial, politeness, affective | Analyzed using LIWC-22 (Linguistic Inquiry and Word Count) dictionary. |
|  | Sentiment | Subjectivity, polarity | Subjectivity score and polarity score, indicating the text's subjectivity and tone. |
| Draft Evaluation Based on Patient Messages | Relevance | Semantic similarity | The semantic similarity between the draft and the patient's message using Sentence-BERT. |
|  | Completeness | Content overlap | Evaluated by the amount of overlap between the draft content and the patient's message. |

To examine the effect of linguistic characteristics on draft utilization, drafts were categorized as either *utilized* or displayed but *non-utilized* by HCPs, and the features of these two groups were compared across the measures described above. This approach allowed us to identify linguistic factors that may facilitate or hinder the adoption of GenAI-generated responses.

#### Effects of utilization on HCP efficiency

Before evaluating the workflow impact of GenAI-draft utilization, we first considered the variability in patient message complexity, as HCPs likely require more time and effort to respond to more complex messages,^72^ potentially masking any time savings or reductions in tasks associated with using GenAI drafts. However, measuring clinical complexity of the message content on a large scale is resource intensive.^73^ To approximate the clinical complexity of messages, we used two proxies: **reading complexity** of messages and **patient complexity** at the time of messaging. Reading complexity was assessed using the same readability measures from **Table 1** and the number of questions in patient messages, quantified by counting sentences that began with wh-words (e.g., why, how, where).^74^ Patient complexity was measured using the Charlson Comorbidity Index (CCI),^75^ which accounts for their age and comorbidities.

To assess the effect of GenAI drafts on efficiency, we adopted a framework (**Table 2**) derived from previous research on EHR-related HCP workload and efficiency.^12,46,76,77^ This framework examined both message- and HCP-level effects. At the message level, we examined whether GenAI draft utilization led to time savings or reduced the number of actions needed to complete a patient message. At the HCP level, we analyzed efficiency by examining the proportion of messages forwarded by clinical and administrative support staff, as physicians rarely forward messages. Forwarding was analyzed only for support staff, assuming messages were intended for the physicians they support per standard team-based workflows. We compared the efficiency metrics between messages with utilized and non-utilized drafts while controlling for patient’s clinical complexity and the draft’s reading complexity. This approach captures both immediate and broader efficiency gains from GenAI-draft utilization.

**Table 2:**
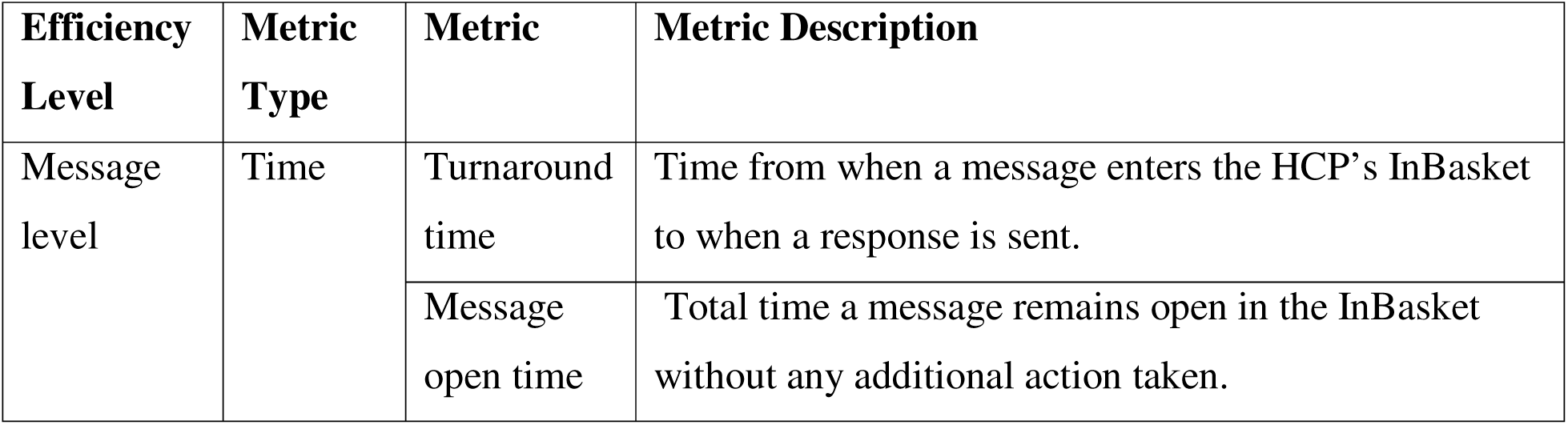
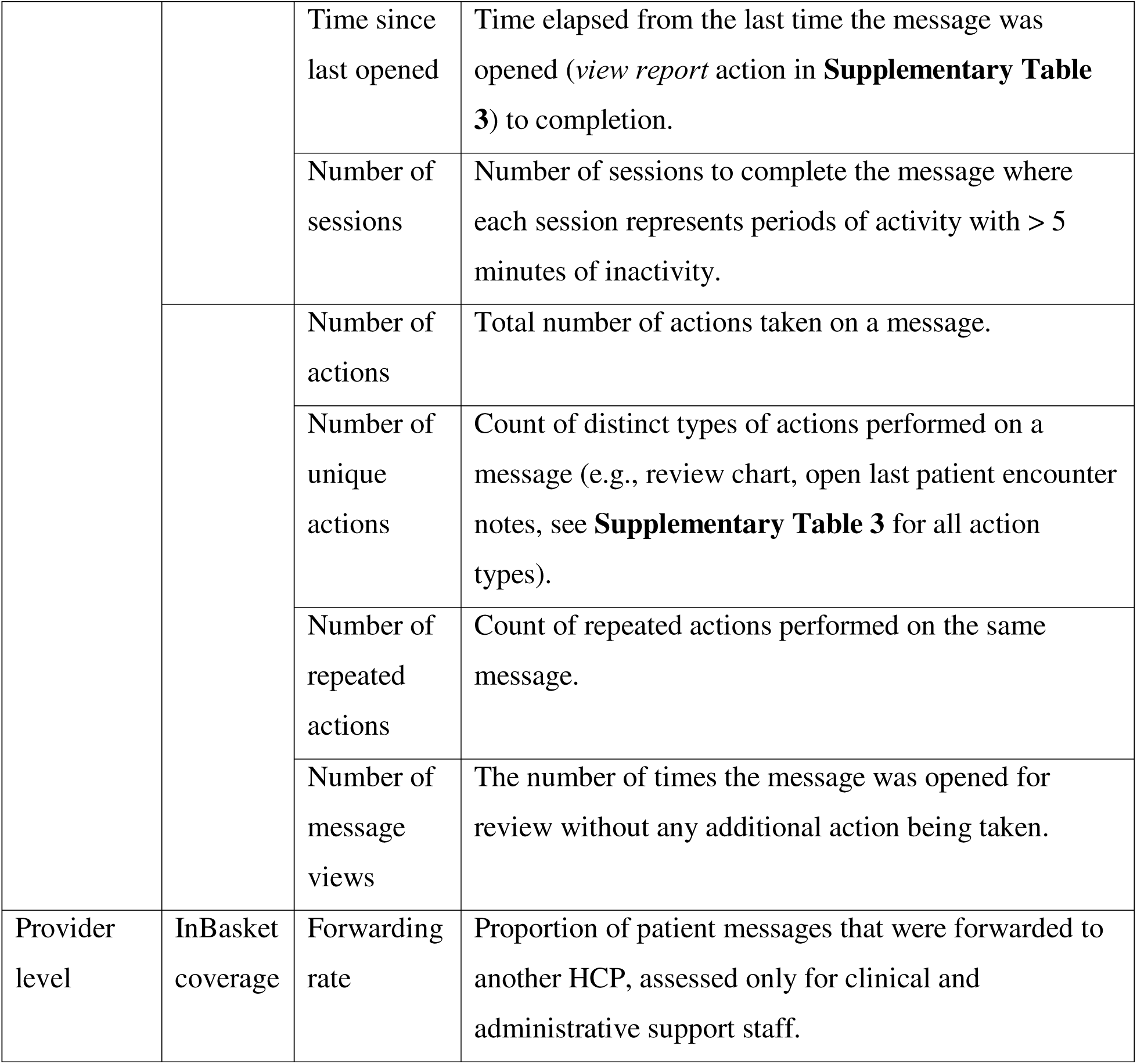
Evaluation framework for measuring the effect of GenAI utilization on HCPs’ efficiency.

#### Data Analysis

Descriptive statistics summarized patient messages, healthcare provider (HCP) actions, and longitudinal utilization trends over two-week intervals, stratified by HCP type. Differences in outcome variables based on GenAI-draft utilization were assessed using univariate analyses at message level. Normality was assessed using the Shapiro–Wilk test, and homogeneity of variance was evaluated using Levene’s test. When assumptions for parametric tests were not met, the Kruskal–Wallis test was used. For significant Kruskal–Wallis results, post hoc pairwise comparisons were conducted using Mann–Whitney U tests with Bonferroni correction to control for multiple comparisons. Categorical variables were compared using the chi-square test. Draft utilization was predicted using a logistic regression model, with HCP type and linguistic features from **Table 2** as predictors, after checking for multicollinearity. Message-level outcomes were analyzed using multivariable regression models adjusting for patient and draft complexity, allowing us to isolate the effect of GenAI-draft utilization on HCP efficiency.

## Declaration Statements

## Data Availability

The primary data on patient messages, HCP actions, and responses used for analysis were sourced from the electronic health record (EHR) data of the New York University (NYU) Langone Health (NYULH) system, containing protected health information (PHI); as such, they cannot be shared publicly. Access to the underlying data requires a Data Use Agreement and IRB approval from NYU Langone Health and its EHR vendor (Epic). While the full content of patient messages and HCP responses cannot be disclosed, de-identified, aggregated metadata on their characteristics can be made available from the authors upon reasonable request.

## Code Availability

The code that supports the statistical analyses and the study findings are available from the corresponding author upon reasonable request. Analysis to process and analyze data was generated with Python v3.11.5.

## Acknowledgements

This work was supported by the National Science Foundation (NSF) grants 1928614 and 2129076 (PI: O.N., Co-PIs: D.M., B.M.W.).

## Author Contributions

S.M., in collaboration with B.M.W., D.M., and O.N., conceptualized the study. S.M. led data collection with support from V.M. and A.C.S., conducted the analyses, drafted the manuscript, and coordinated the coauthors’ feedback. B.M.W. advised on data analysis. W.S., S.R., A.C.S., and D.M. contributed clinical workflow insights critical to data interpretation and results. A.S. provided expertise in implementation science, shaping the Discussion and future directions. B.M.W. and O.N. supervised the project and contributed substantially to writing, revision, and submission. All authors reviewed and approved the final manuscript.

## Abbreviations

EHR: Electronic health record
IB: InBasket
HCP: Healthcare provider
GenAI: Generative artificial intelligence
LLM: Large language model

