## Supplementary Figure 1 for "Utilization of Generative AI-drafted Responses for Managing Patient-Provider Communication"

### Supplementary Information

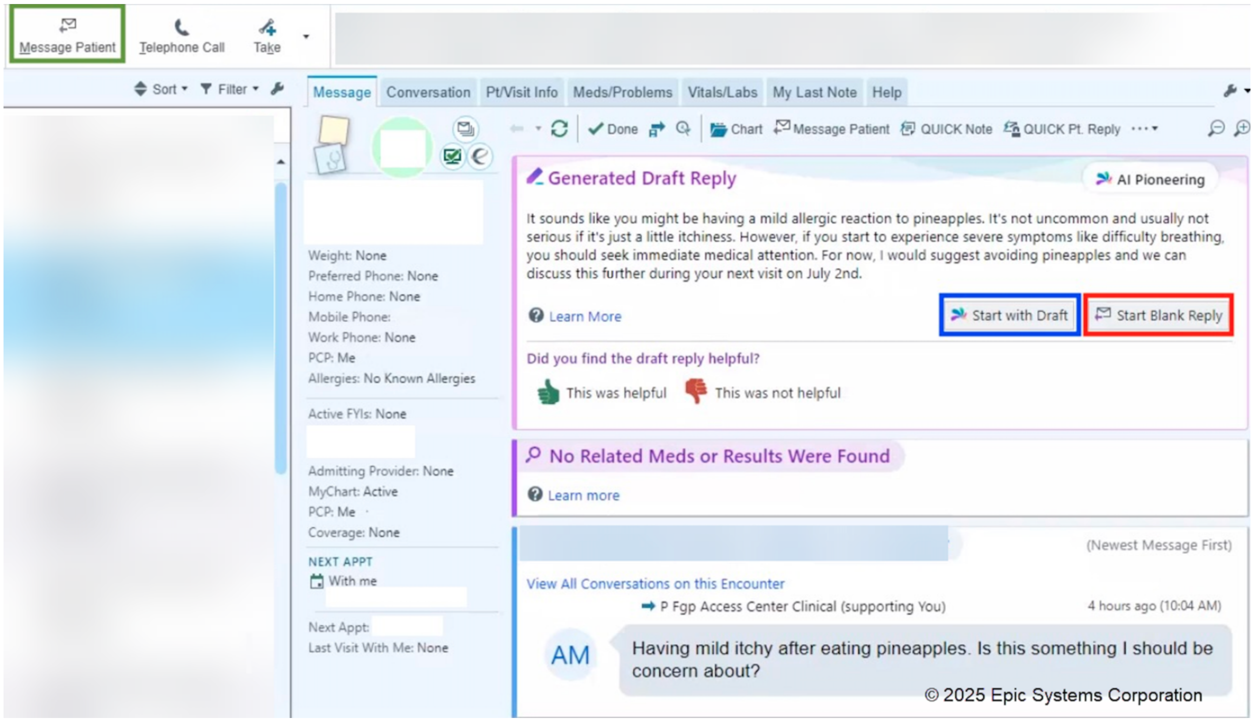


**Supplementary Figure 1**: EHR inbox (Epic InBasket) interface with GenAI augmentation. The three available response options for HCPs are highlighted: **Reply to Patient** (renamed to *Message Patient* in recent EHR update**;** green), **Start with Draft** (blue), and **Start Blank Reply** (red).

| Prompt | Description and Changes | Initial User Feedback on the Drafts |
|---|---|---|
| Baseline prompt | Description:<br>• Original prompt classified patient messages into one of four categories—refills, results, paperwork, or general inquiries—based on message content to guide prompt selection.<br>• Each prompt incorporated the full patient message, relevant EHR data, and contextual details such as the patient's clinical history and | Users noted that the generated drafts tended to be overly lengthy and conversational, which are not consistent with the more concise and professional tone typically used by providers. Many responses often included phrases that invite additional patient messages such as "*don't hesitate to contact me if you have any additional questions*" which raised concerns about additional messaging burden. |

| | | |
|---|---|---|
| | recent/upcoming appointments.<br>• The AI was instructed to act as a clinical assistant, generating responses to patient requests. | |
| Prompt Update 1 | Critical prompt changes:<br>• Do not suggest making an appointment for every request.<br>• Answer the patient messages more directly.<br>• Minimize referencing an upcoming appointment if the appointment is with another provider. | AI drafted responses provided more direct guidance to patients to resolve their issues. More accurate reference of upcoming visits where non-urgent questions could be addressed more thoroughly. Users indicated that the existing message classification had limited usefulness and did not always align with the types of messages providers most frequently receive. |
| Super prompt (current) | Critical prompt changes:<br>• Combined all four categories into a single, unified prompt that specified health system protocols for addressing common questions.<br>• Added additional dynamic elements that auto-populate patient-specific information (*SmartLinks*) and refined prompt for more concise drafts with reduced paraphrasing. | Responses were more aligned with health system operations. |

**Supplementary Table 1**: Summary of prompt structure, key changes, and user feedback that informed the prompt change.

| **Pre-Defined Service Role from EHR (n)** | **Classification (n)** |
|---|---|
| Physician (54) | Physician (54) |
| Medical assistant (6) | Clinical support (14) |
| Registered nurse (4) | |
| Nurse practitioner (2) | |
| Licenses practical nurse (1) | |

| | |
|---|---|
| Technician (1) | |
| Front desk faculty group practice (4) | Administrative support (7) |
| Front desk-2 faculty group practice (2) | |
| ES Manager faculty group practice (1) | |

**Supplementary Table 2: Healthcare professionals (HCPs) included in the study, their roles and classification used for comparing their GenAI-drafted response utilization.**

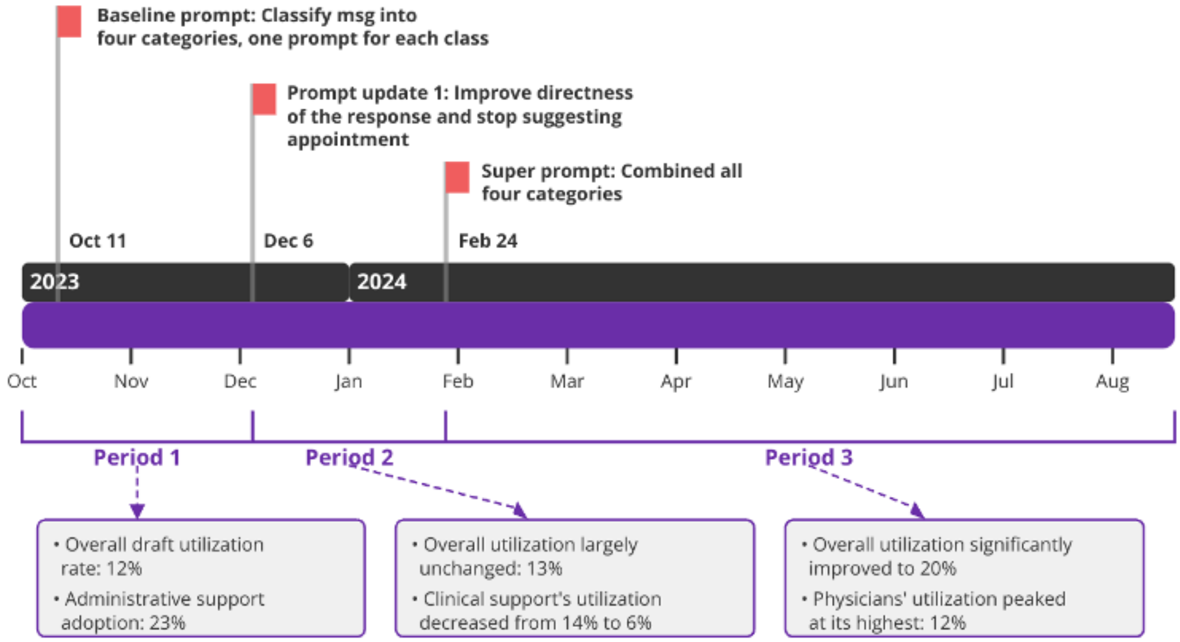


**Supplementary Figure 2**: Prompt updates throughout the study timeline.

#### **Supplementary Note 1**: Inclusion, exclusion criteria

##### Inclusion criteria:

1. Healthcare providers (HCPs) with GenAI-generated drafts enabled in their InBasket.
2. Patient messages received and addressed by these HCPs between October 11, 2023, and August 31, 2024.
3. Patient messages classified as medical advice requests (PMAR) that were written in English and contained no attachments (e.g., images, audio, or video files).

##### Exclusion criteria:

1. HCPs who did not complete the acknowledgment for responsible use of GenAI-generated drafts.

2. Messages where the generated draft was deemed unusable (e.g., draft: "*Unknown*") or failed to load due to system errors (e.g., rate limits, draft length constraints, content filtering, or timeouts).
3. Messages that were not responded to by HCPs participating in the pilot.*,†
4. Messages where the draft was not displayed to the participating HCP, due to technical issues or use of an alternative interface to complete the message where the generated draft could not be displayed.*
5. Messages for which the timestamps of HCP actions in the action log were not available with second-level precision.†

*Additional exclusion criteria for evaluating GenAI draft utilization by the HCPs.
†Additional exclusion criteria when evaluating efficiency in HCP's work due to GenAI draft utilization in time-based measures.

| Sl No. | HCPs' action name | Action description | Count |
|---|---|---|---|
| 1 | View report | Display the selected message | 29540 |
| 2 | Message patient | Reply to patient or send a message using MyChart | 8509 |
| 3 | Patient review | Review chart for this patient | 5802 |
| 4 | Mark as New | Mark the selected message as new | 2327 |
| 5 | Reply to patient (Deprecated- replaced by Message patient) | Reply to the patient using MyChart | 2139 |
| 6 | InBasket suggested response | Reply to patient using Suggested Response | 1434 |
| 7 | InBasket forward message | Forward the message to a new recipient | 700 |
| 8 | Encounter for med review | Open this encounter | 472 |
| 9 | Take responsibility | Mark a message assigned to a pool as yours | 404 |
| 10 | Create telephone call | Create a telephone encounter or open the encounter for this patient. | 361 |
| 11 | Take put back responsibility submenu | Mark a message assigned to a pool as your submenu | 347 |
| 12 | No action required message handled | Mark message as handled and remove from InBasket | 140 |
| 13 | Follow up | View information and take actions on the current message | 76 |
| 14 | Quick encounter note | File a note to the encounter associated with the currently selected message | 67 |
| 15 | Move to my messages | Move selected messages from Completed Work to message recipient's My Messages | 31 |
| 16 | Put responsibility back | Return a message marked as yours to the pool | 15 |
| 17 | Forward message | Forward the message to a new recipient | 14 |

| | | | |
|---|---|---|---|
| 18 | Mark read message | Mark the selected message as read | 11 |
| 19 | Defer to workstation | Defer an InBasket message from mobile device. After deferring, the message will only be available in Hyperspace. | 5 |
| 20 | General properties – no follow-up | View information about the current message | 3 |

**Supplementary Table 3**: Summary of HCPs' InBasket actions and counts for patient messages where GenAI-drafts were generated.

| **HCP Type (n)** | **Utilization** | | | | | | |
|---|---|---|---|---|---|---|---|
| | **Period 1 (11th Oct'23 - 6th Dec'23)** | | **Period 2 (7th Dec'23 - 27th Feb'24)** | | **Period 3 (28th Feb'24- 31st Aug'24)** | | ***p*-value** |
| | **Mean (SD)** | **95% CI** | **Mean (SD)** | **95% CI** | **Mean (SD)** | **95% CI** | |
| Overall (75) | 0.12 (0.13) | [0.07, 0.18] | 0.13 (0.14) | [0.09, 0.18] | 0.20 (0.14) | [0.14, 0.22] | <0.001 |
| Physicians (54) | 0.0 (0) | NA | 0.05 (0.07) | [0.01, 0.09] | 0.12 (0.05) | [0.1, 0.14] | <0.001 |
| Clinical support (14) | 0.14 (0.08) | [0.07, 0.2] | 0.06 (0.07) | [0.01, 0.10] | 0.08 (0.1) | [0.03, 0.12] | 0.143 |
| Admin support (7) | 0.23 (0.41) | [0.13, 0.34] | 0.29 (0.10) | [0.23, 0.36] | 0.32 (0.12) | [0.27, 0.37] | 0.251 |

**Supplementary Table 4**: Comparison of GenAI-drafted response utilization by provider types across different time periods marked by prompt changes. The *p*-values reflect overall groupwise comparisons across the three periods for each provider type, assessed using the Kruskal–Wallis test.

| **Patient message characteristics measure** | **Measure description** | **GenAI-drafted response utilized** | **GenAI-drafted response not utilized** | **Difference in mean** | ***p*-value** |
|---|---|---|---|---|---|
| | | **Mean (SD)** | **Mean (SD)** | | |
| Flesch reading score (0-100 scale) | Measures how easy a message is to read based on word and sentence length (score 70–80: 7th | 76.68 (14.5) | 77.95 (14.74) | -1.27 | 0.005 |

| | | | | | |
|---|---|---|---|---|---|
| | grade level reading) | | | | |
| Lexical diversity (0-1 scale) | Ratio of unique word (stems) to the total number of words (tokens) | 0.79 (0.09) | 0.79 (0.09) | 0 | 0.46 |
| Sentence length per message | Number of sentences per message | 14.7 (6.89) | 14.95 (8.03) | -0.25 | 0.32 |
| Mean syllables per word | Mean in number of syllables within each word in a message | 1.14 (0.13) | 1.12 (0.12) | 0.02 | 0.01 |

**Supplementary Table 5**: Comparison of patient message readability between with and without the utilization of GenAI-draft for responses. The *p*-values reflect differences in distributions between groups, assessed using the Mann–Whitney U test due to non-normality of variables.

| **Evaluation measure (per message)** | **GenAI-drafted response utilized** | | **GenAI-drafted response not utilized** | | **Median Difference** | **Mann-Whitney test stat (*p*-value)** |
|---|---|---|---|---|---|---|
| | Median | IQR | Median | IQR | | |
| Turnaround time (seconds) | 331 | 20,077 | 355 | 13,803 | -24 | 730,738 (<0.001) |
| Message open time (seconds) | 59 | 7,002 | 55 | 4,963 | 4 | 692,073 (<0.001) |
| Time since last opened (seconds) | 23 | 14 | 20 | 65 | 3 | 510,281 (<0.001) |

**Supplementary Table 6**: Analysis of time-based measures of HCP responses to patient Messages: GenAI-draft utilized messages vs. those without it.

| **Evaluation measure (per message)** | **GenAI-drafted response utilized** | | **GenAI-drafted response not utilized** | | **Difference in mean** | **Test stat *(p*-value)** |
|---|---|---|---|---|---|---|
| | Mean (SD) | 95% CI | Mean (SD) | 95% CI | | |
| Number of actions | 4.71 (3.11) | [4.52, 4.89] | 4.62 (3.43) | [4.57, 4.71] | 0.09 | 5,259,935(0.38) |

| | | | | | | |
|---|---|---|---|---|---|---|
| Number of unique actions | 2.82 (0.84) | [2.77, 2.87] | 2.76 (0.8) | [2.74, 2.77] | 0.06 | 5,353,885(0.02) |
| Number of repeated actions | 1.88 (2.73) | [1.72, 2.04] | 1.89 (3.04) | [1.83, 1.95] | -0.01 | 5,165,901 (0.94) |
| Number of message views | 2.61 (2.38) | [2.47, 2.76] | 2.62 (2.62) | [2.57, 2.68] | -0.01 | 5,128,182(0.73) |
| Number of sessions | 2.12 (1.82) | [2.02, 2.23] | 2.06 (1.95) | [2.02, 2.10] | 0.06 | 5,273,79 (0.19) |

**Supplementary Table 7**: Analysis of HCP action counts in responding to patient Messages: GenAI-draft utilized messages vs. those without it.

| **Action Types** | **GenAI-drafted response utilized (n = 1149)** | | **GenAI-drafted response not utilized (n = 10164)** | | **Difference in mean** | ***p*-value** |
|---|---|---|---|---|---|---|
| | Total | Per Message | Total | Per message | | |
| View report | 3019 | 2.63 | 26521 | 2.61 | 0.02 | 0.14 |
| Encounter for med review | 68 | 0.04 | 404 | 0.06 | -0.02 | 0.07 |
| Shift in responsibility (take responsibility, take put back responsibility submenu, move to my messages, and put responsibility back combined) | 37 | 0.03 | 760 | 0.07 | -0.04 | 0.04 |
| Create telephone call | 70 | 0.06 | 291 | 0.03 | 0.03 | 0.03 |

**Supplementary Table 8**: Difference in HCP action types in responding to patient messages between GenAI-draft utilized messages and those without it.

| | **Message forwarded (N)** | |
|---|---|---|
| **Draft utilized (N)** | **False** | **True** |
| **False** | 9565 | 599 |
| **True** | 1079 | 70 |

**Supplementary Table 9**: Contingency table showing the relationship between GenAI draft utilization for responding to patient messages and messages forwarded to another HCP.
